# A qualitative study exploring opportunities for poverty alleviation interventions amongst people with severe mental health conditions in Eastern Cape, South Africa

**DOI:** 10.64898/2026.05.13.26353158

**Authors:** Laura Asher, Bongwekazi Rapiya, Bonginkosi Chiliza, Charlotte Hanlon, Inge Petersen, Carrie Brooke-Sumner

**Affiliations:** Nottingham Centre of Public Health and Epidemiology, School of Medicine, University of Nottingham, Nottingham, UK; Institute of Mental Health, University of Nottingham, UK; Mental Health, Alcohol, Substance Use and Tobacco Research Unit, South African Medical Research Council, Francie Van Zijl Drive, Parow Valley, Cape Town 7501, South Africa; Discipline of Psychiatry, Nelson R Mandela School of Medicine, University of KwaZulu-Natal, Durban, South Africa; Division of Psychiatry, Centre for Clinical Brain Sciences, the University of Edinburgh, Edinburgh, Scotland, UK; Department of Psychiatry and WHO Collaborating Centre in Mental Health Research and Capacity Building, School of Medicine, College of Health Sciences, Addis Ababa University, Addis Ababa, Ethiopia; Centre for Rural Health, College of Health Sciences, University of KwaZulu-Natal, Durban, South Africa; Alan J. Flisher Centre for Public Mental Health, Department of Psychiatry & Mental Health, University of Cape Town, Cape Town, South Africa

## Abstract

**Background:** People with severe mental health conditions (SMHC) and caregivers in South Africa experience high rates of poverty. The PRIZE feasibility trial found that recovery groups were broadly acceptable and feasible and potentially effective in reducing relapse. Addressing economic needs was identified as a means to increase impact. This study aimed to understand experiences of financial insecurity and acceptability of poverty alleviation interventions as an adjunct to psychosocial interventions amongst people with SMHC and caregivers.

**Methods:** We conducted two focus group discussions and 12 in-depth interviews in isiXhosa with a total of 14 people with SMHC and 13 caregivers who had participated in PRIZE in Eastern Cape Province, South Africa. An inductive thematic analysis was conducted.

**Results:** We identified four major themes.

*Theme 1: Financial insecurity as a defining influence on life:* We found that financial security was crucial to recovery, through bringing status and dignity. However, participants experienced substantial financial insecurity, which impacted on social and mental wellbeing. Financial insecurity was entrenched due to fractured and violent communities, cycles of debt and stigma amongst employers.

*Theme 2: Government disability grants are not a panacea.:* Difficulties accessing disability grants included problems attending assessments and rejection of applications. Whilst they were generally welcomed, receipt of disability grants sometimes caused problems such as increased stigma and family disagreements about how the money should be spent.

*Theme 3: Group savings offer conditional hope if carefully managed.:* Several caregivers had longstanding experiences of stokvels (community-based credit unions). However, some were fearful of group members absconding with funds. Participants emphasised that trust, safety and fairness are essential for successful group savings.

*Theme 4: Income-generating activities are desired but need capital and come with safety concerns:* Many had ideas and motivation for small businesses but stressed the need for financial capital, skills training and financial literacy support. There were serious concerns that owning a business or gaining wealth could make one a target of crime.

**Conclusion:** Poverty alleviation interventions could positively impact on the wellbeing of people with SMHC and caregivers in South Africa as an adjunct to psychosocial interventions and psychiatric care. Approaches could include supporting access to social protection or existing savings groups, and nesting new savings groups or income generation initiatives into psychosocial interventions. Any model would need to incorporate robust mechanisms to ensure the safety of participants. All approaches would be enhanced by parallel social and public health interventions to build social capital and reduce violence in neighbourhoods.

## Introduction

People with severe mental health conditions (SMHC) are more likely to live in poverty than the general population in the Global South (Tirfessa et al., 2017). Disabling symptoms, medication side-effects and discriminatory exclusion are barriers to employment and other economic activity (Asher et al., 2023a). Unemployment amongst people with SMHC in South Africa is around 93% (Wootton et al., 2024, Asher et al., 2024), compared to 19.6-40.4% in the general population (Statistics South Africa, 2025). Increased health expenditure, and the sacrificing of work to care for relatives, further impact household financial security (Rapiya et al., 2025, Dijkstra et al., 2025). Lack of participation in the world of work accelerates social exclusion and may be one pathway to severe forms of social marginalisation such as homelessness (Hanlon et al., 2025). Epidemiological evidence also suggests economic inactivity accounts for a substantial proportion of mortality amongst people with SMHC (Das-Munshi et al., 2025). Moreover, poverty acts as a critical barrier to accessing mental health care, particularly in settings where treatment for severe mental health conditions is not included in universal health coverage (Mlay et al., 2025, Mlay et al., 2022). Poverty alleviation interventions are therefore a priority amongst people with SMHC in South Africa (Brooke-Sumner et al., 2024b) and other countries in the Global South (Asher et al., 2018).

Various types of poverty alleviation interventions are relevant for people with SMHC, and suitable approaches may vary depending on the social and economic context. First, social protection measures for people with disabilities, including people with SMHC, are available in much of the Global North (Stevelink et al., 2024). In South Africa disability grants of R2092/£90 per month are available for individuals whose illness impairs their ability to work (assessed via a functional assessment completed by a medical doctor), as long as they also satisfy a means test (yearly earnings of less than R86,282/£3,695 in 2023), and are not in the care of the state (Wootton et al., 2024). In practice individuals may also be excluded from accessing the disability grant if they are not engaged in psychiatric care. Second, interventions to support access to social benefits or external livelihoods schemes (for example, through signposting) may be provided as an adjunct to broader psychosocial interventions. The approach has been taken in research studies (Chatterjee et al., 2014) and real-world programmes (Shields-Zeeman et al., 2017) in India as well as being one aspect of peer support groups in Kenya (WHO, 2021b). Third, livelihoods interventions, such as savings groups or microfinance initiatives, can be directly embedded within psychosocial interventions supporting people with SMHC, typically delivered alongside provision of mental healthcare. These approaches tend to be used by non-governmental organisations (NGOs), such as Basic Needs (Raja et al., 2012, de Menil et al., 2015).

There is a clear policy directive to address economic concerns amongst people with SMHC within South Africa and globally. The World Health Organisation Mental Health Action Plan 2013–2030 highlights the importance of financial protection for socioeconomically disadvantaged groups (WHO, 2021a). The South African National Mental Health Policy commits to addressing the ‘poverty-mental health link’ though inclusion of people with MHCs in education, skills development and income generation opportunities, social protection, and workplace accommodations (Department of Health, 2024). However, real world implementation of these commitments is challenging and slow.

In 2019-2023 our team conducted the Peer-led Recovery Groups for People with Psychosis in South Africa (PRIZE) randomised controlled feasibility trial (Asher et al., 2024, Asher et al., 2023b, Brooke-Sumner et al., 2024b, Brooke-Sumner et al., 2024a). Recovery groups were held weekly in community settings with groups of people living with psychosis and their caregivers. The groups used a strength-based, recovery-oriented approach comprising check-in, discussion of key topics (such as understanding mental health, healthy relationships, setting goals) guided by stories, group problem-solving, and socialising. We found that recovery groups are acceptable, feasible and may be effective in reducing relapse amongst people with SMHC in an underserved setting in Eastern Cape (Asher et al., 2024, Asher et al., 2025). However, participants suggested that groups would provide further benefits by including a poverty alleviation component, which was also highlighted as a need in previous South African recovery-focused work (Brooke-Sumner et al., 2017). Research in other Global South settings has found that inability to address poverty may be a barrier both to impact of psychosocial interventions (for example, if participants cannot afford mental healthcare) and acceptability (that is, an unwillingness to engage in an intervention that does not address their most pressing needs)(Asher et al., 2018).

A co-operative is an organisation democratically owned and controlled by its members. There is a long history of co-operatives in the African continent, particularly those focused on agriculture and inclusive development. A stokvel is the name given to South African community-based co-operative savings group where members contribute a fixed amount of money into a shared pool at regular intervals (usually weekly or monthly). Stokvels are a common approach to saving in South Africa, with an estimated 800,000 groups and 11 million members nationwide. Stokvel group members use the communal fund for a predefined purpose such as savings, funeral expenses or grocery purchases (National Stokvel Association of South Africa, 2025). Savings groups are also a widespread approach in the NGO/ development sector in South Africa, often targeting women and other vulnerable groups as a means of building sustainable livelihoods (SAVEact, 2026). However, to our knowledge these endeavours do not typically target people with SMHCs as a vulnerable group. This study aimed to understand experiences of poverty and current engagement with poverty alleviation interventions, including disability grants for people with SMHC and their caregivers in Eastern Cape, South Africa. We also aimed to interrogate potential poverty alleviation components for incorporation into future psychosocial interventions, including stokvels as a contextually appropriate model.

## Methods

### Setting

The study was conducted in low-resource urban and semi-urban areas (known as townships) of Nelson Mandela Bay Metropolitan Municipality, Eastern Cape Province, South Africa. Townships are the legacy of apartheid era planning strategies that forcibly relocated Black residents to the urban peripheries (Matzopoulos et al., 2020). These areas have a high concentration of insecure housing, limited infrastructure, high rates of alcohol and drug abuse and community violence (Christie et al., 2020). The unemployment rate in Eastern Cape is 39.3%, which is the second highest rate by province nationally (Statistics South Africa, 2025). The province also has the highest murder rates nationally at 15.4/100,000 population (South African Police Service, 2025). In Nelson Mandela Bay district 94% of the population has access to flush toilets and 98% has access to electricity. The majority of the population is Black African (60%) or ‘Coloured’ (mixed ancestry) (24%) (Nelson Mandela Bay Municipality, 2020).

The district has eight secondary level hospitals and one specialist psychiatric hospital. There are 41 primary health care (PHC) clinics, eight of which provide mental health services delivered by psychiatric nurses, focusing on repeat psychotropic medication, which is free but not always available. Aside from PRIZE, there is no community-based provision of psychosocial support for people with SMHC.

### Participants

PRIZE participants were originally identified through PHC clinics then were randomly allocated to recovery groups plus treatment as usual (TAU) or TAU alone. A subset of people living with SMHC (n=14) and caregiver (n=13) participants were selected purposively for this study by age, gender, clinic and trial arm. Of these there were nine linked people living with SMHC - caregiver pairs (i.e. 18 of 27 participants). The inclusion criteria were: (i) experience of SMHC or caregiver to someone with SMHC, (ii) over 18 years (iii) person with experience of SMHC having capacity to give informed consent (assessed using a tool developed in PRIZE). Potential participants were contacted by phone in person at recovery group sessions. They were given written and verbal information and written informed consent was obtained. Recruitment was carried out between 1^st^ -31^st^ October 2023. No potential participants refused or dropped out.

### Data collection

We initially conducted two focus group discussions, with participants divided into those who had participated in recovery groups and those who had not. Both groups included a mixture of people living with SMHCs and caregivers. FGDs covered experiences of financial insecurity, disability grants, and savings groups and the acceptability of group savings models. We then conducted 12 in-depth interviews (7 people living with SMHCs and 5 caregivers) to explore emerging topics and individual experiences in greater depth. This also gave people living with SMHC and caregivers an opportunity to discuss topics that may be sensitive to discuss in the presence of their relative. There was no one else present besides the interviewer and participant. Data saturation was considered reached when consecutive interviews yielded no novel thematic content. FGDs and IDIs were conducted in isiXhosa using an interview guide. FGDs and IDIs were conducted by BR (MSc), who is a female first language isiXhosa speaker and experienced qualitative interviewer and research coordinator. She had had previous contact with recovery group participants through observations made as part of the PRIZE process evaluation. The FGDs and IDIs were conducted in person in a private space in the participant’s home or at a local venue, or by telephone. Caregiver IDIs lasted between 26 and 44 minutes, people living with SMHC IDIs lasted between and caregiver IDIs lasted between 12 and 53 minutes and FGDs lasted 76 and 97 minutes. FGDs and IDIs were audio-recorded, transcribed and translated to English by BR. Participants received modest remuneration (150 Rand) for their time and transport costs.

### Analysis

We used inductive thematic analysis (Braun and Clarke, 2006), paying particular attention to the conceptualisation of themes as ‘creative and interpretative stories about the data’, as opposed to domain summaries (Braun and Clarke, 2019). We also sought to apply a socio-political determinants lens to the data (Burgess et al., 2025). The analysis was conducted in Microsoft Word. CBS and LA first familiarized themselves with the transcripts. CBS identified an initial coding framework and constructed themes and sub-themes from across the data by grouping conceptually similar codes. LA amended themes iteratively and collaboratively through discussion with CBS and coding of further manuscripts. The study is presented in line with COREQ guidance for reporting qualitative research (Tong et al., 2007) (Supplementary file 1). The South African Medical Research Council Ethics committee (EC027-6/2021) gave ethical approval for the study.

## Results

We included 14 participants living with SMHC and 13 caregiver participants in the IDIs and FGDs (Table 1). All participants living with SMHC, except one, were male; whereas all caregiver participants were female. All participants were Black African. The majority of participants living with SMHC were unemployed (86%) and in receipt of the disability grant (79%). Four major themes emerged: 1- Financial insecurity as a defining and intractable influence on life; 2- Government disability grants are not a panacea, 3-Group savings offer conditional hope if carefully managed, and 4- Income-generating activities are desired but need capital and come with safety concerns.

**Table 1.** Participant characteristics.

|  | Characteristic | N (%) |  |
| --- | --- | --- | --- |
|  |  | People living with SMHC | Caregivers |
| **Gender** | **Male** | 13 (93%) | 0 |
|  | **Female** | 1 (7%) | 13 (100%) |
| **Age** | **20-39 years** | 5 (36%) | 3 (23%) |
|  | **40-59 years** | 7 (50%) | 5 (39%) |
|  | **60 years or more** | 2 (14%) | 3 (23%) |
|  | **Missing** | 0 | 2 (15%) |
| **Race** | **Black African** | 14 (100%) | 13 (100%) |
| **Employment status** | **Unemployed** | 12 (86%) | 6 (46%) |
|  | **Employed or self-employed** | 2 (14%) | 2 (15%) |
|  | **Pensioner** | 0 | 4 (31%) |
|  | **Missing** | 0 | 1 (8%) |
| **Highest level of education** | **Primary education only** | 2 (14%) | 2 (15%) |
|  | **Secondary education** | 11 (79%) | 9 (70%) |
|  | **Higher education** | 1 (7%) | 0 |
|  | **Missing** | 0 | 2 (15%) |
| **Access to disability grant** | **Disability grant** | 11 (79%) | - |
|  | **No disability grant** | 3 (21%) | - |
|  | **Total** | 14 | 13 |

### Theme 1: Financial insecurity as a defining and intractable influence on life

Financial security was crucial to recovery, but participants experienced substantial financial insecurity, which impacted on wellbeing. Financial insecurity was entrenched due to fractured and violent communities, cycles of debt and stigma amongst employers.

#### Subtheme 1a: Financial security as crucial to recovery

Financial security seemed to offer various routes to personal recovery through bringing peace of mind, dignity, motivation and the ability to make and action plans. For some people living with SMHC, visibility of wealth was important- dressing well and looking good meant feeling good too. One caregiver felt that social contact opportunities gained through employment would improve their relative’s illness. For several participants, money would allow individuals to fulfil valued social roles such as having and providing for children, or contributing to Xhosa rituals. Financial independence and the ability to help others was felt to bring status in families and communities.

> *“my family would want me to help where I can help and also, they will be proud that I am independent. If they want to be helped with something, I will help them. These kids need to be provided for and not to be forgotten…. This would be a big challenge if there is no money, that is how I’m thinking about it. I think they would be proud because when there are things that needs to be done at home like maybe there is a ceremony, you are able to help. Things like that. Do you see that you’re growing, and they will be proud of you”*
>
> (Male living with SMHC IDI14)

### Subtheme 1b: Financial insecurity impacts social and mental wellbeing

Caregivers and people living with SMHC described multiple competing demands on finances, including costs related to children living in their home. Many participants spoke of having insufficient funds to meet their needs. After paying for basics such as bills and groceries, there was often nothing left for other essentials, such as children’s shoes. A minority of individuals reported skipping meals or avoiding expensive food such as meat. These financial stressors caused tangible impacts on mental health, including suicidal thoughts in some people living with SMHC. In some cases, poverty led to alcohol and substance use, which in turn worsened family and social relationships. Family conflict also resulted from the unemployment of some people living with SMHC and the caregivers’ tendency to shoulder the responsibility for the household functioning including finances. The broader impact of widespread poverty was acknowledged, with violence in communities reportedly exacerbated by hunger, and discord between community members traced back to financial inequalities.

> *“When we are running low or have run out of things in the house or there is shortage somewhere, [my son] becomes really stressed we even argue about irrelevant stuff. So, I have come to understand that stress is the reason behind our arguments because there is no money, and the fact that he is unemployed is making things worse, things would have been better if he was working”*
>
> (Female caregiver ID57)

Several participants spoke of needing money to contribute to traditional Xhosa ceremonies, such as male circumcision, or to attend church. These ceremonies are highly valued in Xhosa society, and the lack of funds and impacts on participation was a source of stress and sadness.

#### Subtheme 1c: Fractured communities entrench individuals in financial insecurity

Whilst a small minority reported being able to save money, the vast majority found it impossible to do so. Reliance on loan sharks (informal lenders in the community charging high interest rates) meant some individuals were “*drowning in debt*” *(Female caregiver IDI57)*. Moreover, some caregivers complained that their relative spent money recklessly on alcohol rather than spending on essential needs, which was seen as a key route to entrenchment in poverty. Participants reported extremely limited opportunities for education, skills training and employment in the townships where they lived. Getting public transport to locations with better job-seeking opportunities was expensive and potentially fruitless. Some reported the few opportunities locally were unobtainable due to nepotism or unavailable to Black residents.

> ***“****I went to the councillor the other day to ask if I could cut the grass in the park, but such kind of jobs are never given to you as Black residents of that community, instead you will see these parks being cleaned and grass cut by other people from other areas that are White. So, it is difficult for us to get positions.”* (Female caregiver, TAU FGD)

Difficulties finding work that could accommodate periods of mental ill health, physical disabilities or medication side effects, such as dizziness, were cited by some participants. For others, stigma amongst employers was felt to be a key barrier to employment.

> *“we cannot work in restaurants because they say we are dirty…You know even in TV they laugh at mental illness; they make fun of it. For instance, if I want to become a nurse, I won’t be able to get a job in the hospital because they will say I will kill the patients. You get undermined in all aspects; they don’t understand this illness because it is not happening to them.”*
>
> (Male living with SMHC IDI57)

Whilst many caregivers supported their relative financially and, in some cases, tried to protect them from loan sharks, in many cases financial support from the extended family or community was absent. This was due to high levels of poverty amongst all in these neighbourhoods, mental illness stigma or a general lack of solidarity. Those who had attempted small businesses (for example, selling sweets or electrical cables, or sewing) were frequently thwarted by the inability of customers to pay for produce.

> *“I sell things but my business has not been successful because you wish you could get some support. I had a sewing business, but you will find that people do not pay and that would mean that you will need to take it from your pocket for you to be able to survive. So, I stopped because if I do, it’s difficult to get people to pay.”*
>
> (Female caregiver, TAU FGD)

The high prevalence of violent crime characterizing the areas where participants live magnified the impact of poverty. In one instance, a caregiver was receiving financial support from her son until he was shot dead. Fear of robbery was cited as holding people back from acquiring possessions.

> *“on one side, you do wish for something, and you feel you can be able to get this thing for yourself. There is that fear, for example, I would wish to get a plasma TV, I would work to put together the money for it. You don’t know how long it took me to put this money together and how. I will then buy this plasma. Within three months my door is kicked, and this plasma is taken. So, we live in such fear. You will find that you were able to put money together but now you are scared of a person. You understand?…so, we are held back.”* (Female living with SMHC, recovery group FGD)

### Theme 2: Government disability grants are not a panacea

There were difficulties accessing disability grants, and whilst they were generally welcomed, receipt of disability grants sometimes caused problems within families.

#### Subtheme 2a: Difficulties in accessing disability grants

Whilst participants welcomed the provision of disability grants there were manifold difficulties in accessing them. One caregiver had initially not been informed by doctors that their relative would be eligible, so she did not apply. In some cases, there were practical barriers to attending assessments, including the long distance from home, long waiting times, and technical problems resulting in cancelled appointments. Others described being rejected due to being incorrectly assessed as fit for work, or due to alcohol consumption being picked up on a urine test. In some cases, these difficulties spanned several years before the grant was obtained.

> *“You see, I received [the disability grant] after 10 years, no, after 15 years of trying, I was rejected based on the fact that I was told I am fit for work….I was fit for work, and at that time I didn’t have clothes to wear, I wore torn and old clothes, I was extremely stressed to the extent that it felt as if I was not taking my treatment”.*
>
> (Male living with SMHC IDI57)

Whilst some participants reported no problems with receiving the grant once it had been accessed, in other instances the grant was suspended for several months because the individual disengaged from care, or conversely was receiving inpatient treatment, or whilst the application was being renewed. One individual had to repeat the application process due to moving province. Two people living with SMHC reported a several month gap in receiving the grant during the Covid pandemic.

#### Subtheme 2b: Receipt of disability grants comes with challenges

In many families disability grants seemed to be pooled with other household income and spent on basic communal needs such as food. Less commonly the grant was seen as income for personal use. Whilst one person living with SMHC spoke of being able to buy important items, including a bed and fridge, most participants felt disability grants were insufficient to support all basic needs let alone further their economic security. No participants reported using their disability grant to save, join a stokvel or start a small business. Moreover, receiving a grant created various difficulties in participants’ lives.

For some people living with SMHC, receiving the grant was a sign of sickness and therefore a source of embarrassment and loss of dignity which caused them to be mocked by family and others.

> *“P (Participant): I am tired of being teased about the social grant.*
>
> *I (Interviewer): Who is teasing you? Your friends?*
>
> *P: I am teased at home and everywhere.*
>
> *I: What are they saying?*
>
> *P: They say I am being paid a social grant. A person who is paid a social grant is someone who is sick.*
>
> *I: Mm. But even old people get paid a social grant even though they are not sick. How does this make you feel?*
>
> *P: I don’t feel great.*
>
> *I: Ok. I hear you.” (Male living with SMHC IDI91)*

In some cases, grant money caused arguments within households about how it would be spent. One man living with SMHC reported having to fend off requests from acquaintances to share the money; they felt this was fair game as the individual had not worked for it. In other instances, when money was entirely controlled by caregivers this lack of agency was painful and insulting for people living with SMHC.

> *“P (Participant): no, sister even for us people who have mental conditions money does cause problems. When you get paid a person would keep your money and say “leave it I will keep it for you”*
>
> *I (Interviewer): how does that make you feel?*
>
> *P: it does not sit well with me, it hurts me, because it feels like you are taken as a person who doesn’t have a brain and yet you go for your treatment and taking your pills and you are OK but when payday comes you have to take your card and give it to them to get the grant money for you*.
>
> *When you ask for the card, you are asked “what are you going to do with it?””*
>
> (male living with SMHC, recovery group FGD)

Some participants reported that receiving the grant did not impact on ability or willingness to seek employment. Some said the ideal situation was to have both, whilst others would prioritise employment due to greater dignity and better income. Yet there was confusion amongst other participants as to the maximum wage allowed whilst receiving the grant, or whether employment was permitted at all. One caregiver described how her relative/son had been turned away from job opportunities due to receiving the grant.

### Theme 3: Group savings offer tentative hope if carefully managed

Several caregivers had longstanding involvement in stokvels but there were some negative experiences and expectations. Participants emphasised that trust, safety and fairness are essential for successful group savings.

#### Theme 3a: Mixed experiences of stokvels

Many participants, mainly caregivers, had been members of stokvels at some time, with one caregiver reporting membership for 20 years, and the approach was familiar even to those who had not participated. Some spoke of their success in saving and purchasing household items such as a washing machine, tiles, couches and TVs. Most participants saw the value in saving and said they would join a savings group if it would help them to achieve this. Some cited the reduced reliance on loan sharks therefore minimizing debt, and in other cases the emphasis was on the development and empowerment participation in a stokvel offered.

> *“This [group savings] would give you an opportunity to show people who think that when you have money you don’t know how to use it that you can. If this idea could happen, even your walk would change. You feel that you have been born anew, you are Nicodemus.”* (male living with SMHC, TAU FGD)

However, difficulties with stokvels were also reported. Some felt they would not be able to participate- or had quit a group- because they did not have enough money contribute.

> *“I (Interviewer): is [a savings group] something that you would join?*
>
> *P (Participant): I have a problem; I have a lot of debt.*
>
> *I: You wouldn’t be able to join because you have a lot of debt?*
>
> *P: the money that I get is not enough, I can [join] if only I have a lot of money or I have won lotto, something like that.”* (male living with SMHC, recovery group FGD)

One caregiver said they would not have time and another was already of part of a stokvel so would be wary of overcommitting by joining a new group. There were also major concerns that group members would not contribute or not pay back money, leading to fighting and financial losses (“*they eat people’s money in these stokvel*s.” (Male living with SMHC IDI26)). Some had experienced this through previous groups, whilst others had heard such stories on the radio or through friends. For some this would result in creating enemies amongst people they knew. There was also significant anxiety- sometimes due to personal experience- that due to the pooling of cash stokvel members could be targets for armed robbery.

> *“[they] fetched the money and went to one of the stokvel members’ house. When they got there, there was a car waiting for them. They did not even get into the house. They got robbed outside. A gun was pointed at them and that money that was fetched amounted to some hundreds of thousands was taken. Right in front of the gate!” (female caregiver, TAU FGD)*

Alternative ways of saving were mentioned, including funeral associations and ShopRite grocery store stamps (whereby shoppers buy physical stamps at the till, paste into a savings booklet and then typically use the stamps to buy groceries store cards at high-expense times, such as the December holiday season).

#### Subtheme 3b: Trust, safety and fairness are essential for successful group savings

Provisions to ensure fairness and safety of funds were seen as crucial. It was emphasised that group members should be honest and able to trust each other. As such it was seen as essential for participants to know one another; PRIZE groups were seen by some as a potential starting point. One participant mentioned that being a permanent resident in the area should be an entry criterion.

> *“you need to make sure that it’s people who are trustworthy. You can’t just allow anyone to join because you may contribute towards that person when it’s their turn but when it comes to you, they don’t do the same. You must trust each other and also know each other.”* (female caregiver, recovery group FGD)

Participants highlighted that some caregivers control the money of people living with SMHC, and that people living with SMHC had complained about this in PRIZE sessions. It was felt that for savings groups people with SMHC should have financial autonomy, and not be coerced into participation, or have their funds controlled, by caregivers. Linking savings groups to PRIZE, and having funds dispersed by social workers, was felt to give credibility and security to money sharing arrangements. Finally, the importance of setting realistic savings targets was raised given the competing priorities on participants available income.

### Theme 4: Income-generating activities are desired but need capital and come with safety concerns

Many had ideas and motivation for small businesses but stressed the need for financial capital, skills training and financial literacy support. There were serious concerns that owning a business or gaining wealth could make one a targets of crime.

#### Subtheme 4a: Need for financial capital, skills training and financial literacy support

Many had the ideas and motivation for diverse microenterprises, such as woodworking, haircutting, grass cutting, drawing, growing and selling food. These proposals had sometimes been discussed at PRIZE groups, but in most cases planned activities didn’t materialize. There was a sense that whilst such initiatives could conceivably be embedded in PRIZE groups, substantial support was needed for ideas to become a reality. Some participants emphasised the need for skills training, whereas others stressed that they already had the required skills, such as mending shoes and carpentry, but lacked space in which to carry out the work. A few participants also saw support with financial literacy as important, for example advice on how to avoid debt.

Moreover, a major barrier was the lack of capital; this was needed to buy tools, petrol, seedlings, beads or paint.

> *“So, even if you wish to do start something, you’d realize that you don’t even have a capital.” (female caregiver, recovery group FGD)*

#### Subtheme 4b: Neighbourhood crime thwarts income-generation efforts

There was often strong desire to engage in income-generation activities but again neighbourhood violence had a powerful impact on the types of opportunities that were perceived to be viable. Small business owners were seen as particular targets for violence and theft, with some recounting stories of fatal shootings. In addition to concerns about holding quantities of cash (subtheme 3b), one participant described having to pay protection money to guard against robbery, which was a substantial dent on their earnings from selling fish.

> *“another thing that prevents people from having businesses in the community it’s because you would start a business and then the “amaphara” [thief and/or person who uses drugs] will come and they will take everything that you have brought causing you to be unable to start your business.”*
>
> (Female caregiver, TAU FGD)

These issues were compounded by the challenge of finding local customers with sufficient resources to buy the produce or service being offered (subtheme 1a).

## Discussion

This study has presented in-depth findings on the experiences of poverty affecting people with SMHC and their families in South Africa. In our study, mental health and poverty were connected in a bidirectional relationship that affected both people with SMHC and their caregivers (Lund et al., 2011). Whilst mental health problems led to exclusion from employment, financial insecurity was itself a major source of stress and a barrier to personal recovery. Conversely, financial security was felt to be a route to recovery through increased social inclusion and self-esteem. This reflects findings from other African settings that work and economic prosperity are both an outcome and a process of mental health recovery (Asher et al., 2026, Vera San Juan et al., 2021). Our participants painted a picture of fractured, unstable, violent, and isolated communities. The shadow of crime and violence seemed to seep into multiple aspects of why people become and stay poor: from the murder of family members, to the fear that gaining assets or starting a business would make one a target, to the perceived untrustworthiness of fellow stokvel members. These findings highlight the complexity of bringing individuals and families out of entrenched poverty in South African township settings. Further they suggest that participating in group savings or income-generating activities could actually increase fractures, by creating enemies amongst existing social networks. All these experiences were amplified by the presence of mental health conditions: it was harder to get a job, there was less support available and even more complications in doing group savings. These are key considerations for meaningful and appropriate poverty alleviation interventions in this setting.

These proximal factors were in turn underpinned by broader socio-political determinants (Burgess et al., 2025). The legacy of apartheid manifested in the spatial segregation of townships from employment opportunities and transport links (Luke 2025), as well as in direct experiences of racial discrimination. Despite strenuous legislative and policy efforts to eliminate racial inequalities in employment and economic status (Busse 2025), substantial inequities still exist in South Africa.

### Implications

As mental health and financial security are so tightly entwined it is arguably imperative to address the economic situation of people with SMHC alongside other aspects of treatment services. However, the most effective, feasible, acceptable and ethical way to support economic needs is unclear. Our findings suggest that intervening in the economic ecosystem of people with SMHC and their families could have serious consequences, potentially causing conflict within families and communities, and even threatening the safety of participants. There have been minimal evaluations of poverty alleviation models for people with SMHC (de Menil et al., 2015, Mlay et al., 2025) either within or outside of psychosocial interventions (Asher et al., 2017). A recent systematic review examining impacts of poverty-reduction interventions combined with psychological interventions did not identify any studies targeting people with SMHC (Tanski et al., 2025).

Psychosocial interventions for people with SMHC that have tried to address poverty indirectly, for example through community-based rehabilitation targeting a return to farming work, are effective in improving functioning but not in improving economic outcomes (Asher et al., 2022, Asher et al., 2023a). There is some evidence from low-income countries that expanding access to mental health care may improve economic status and food insecurity by reducing catastrophic out of pocket expenditure (Tirfessa et al., 2020, Hailemichael et al., 2025).

Based on our results, we have identified four potential approaches to providing a poverty alleviation component as an adjunct to psychiatric care and psychosocial interventions, such as PRIZE recovery groups, that would be suitable for low-resource settings. Each differs with respect to (i) potential impact on financial insecurity and/or mental health, (ii) scalability and sustainability, and (iii) safety and ethical concerns (see figure 1).

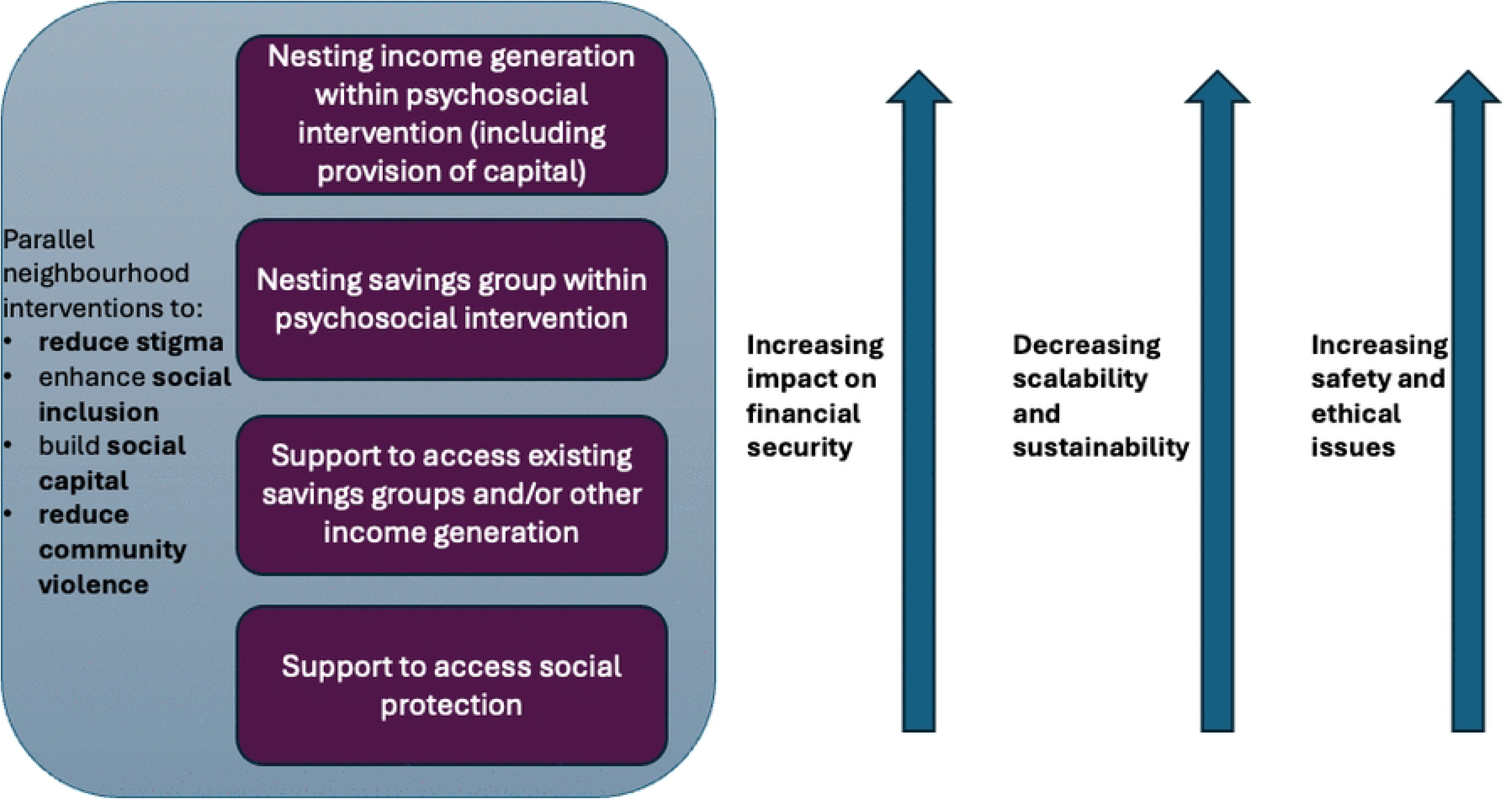

#### 1. Support to access social protection

Our study found that disability grants were welcomed, and allowed some needs to be met. This reflects a large South African cross-sectional survey (n=1154) which demonstrated that receipt of disability benefits is significantly associated with increased personal and household wealth amongst people with SMHC (Wootton et al., 2024). Qualitative evidence suggests unconditional cash transfers for people psychosis in South Africa delivered as part of a research project were typically used for transportation to hospital and buying food. There also appeared to be indirect positive impacts on family relationships, medication adherence and stress amongst people with psychosis and caregivers (Mlay et al., 2025).

The difficulties we identified with accessing disability grants have been highlighted by the UN special rapporteur on extreme poverty and human rights (United Nations General Assembly, 2024). Addressing these barriers is an area of focus in the South African Mental Health Strategy, with planned approaches including development of guidelines to facilitate access to social grants amongst people with mental or intellectual disabilities and development of supportive arrangements for continuation of grant support during periods of review and transition (Department of Health, 2024). Mental health NGOs could have an important role in supporting this policy direction, for example clarifying eligibility and helping individuals to navigate the system. Auxiliary social workers successfully undertook this role in the PRIZE trial (Asher et al., 2024). There is a growing literature indicating that cash transfers may be associated with modest improvements in mental health (McGuire et al., 2022) and positive economic outcomes (Tanski et al., 2025). However existing evidence is focused on the general population or people with common mental disorders. With some exceptions (Mlay et al., 2025, Mlay et al., 2022) there is little previous research on the direct effects of, or the signposting to, social protection amongst people with SMHC in the Global South. Real-world programmes using these approaches, including the Basic Needs Model, have not been formally evaluated for this population (Pathare et al., 2023, WHO, 2021b).

In our study we found that disability grants tended to be pooled with other household income. Whilst this may be a typical, culturally appropriate practice, there were signs that people with SMHC were considered to have less say in how communal funds were spent compared to other family members. In some cases SMHC found this distressing and/or wanted more control, or family conflict was caused. In other cases people living with SMHC felt shame around receiving a disability grant. To address these issues, the role of ASWs could be developed to support the psychological and relational impacts of receiving benefits. Alongside this issues around interpersonal power dynamics, rights and autonomy of people living with SMHC would need to be explicitly explored and negotiated. The PRIZE format – which involved people living with SMHC and caregivers working both separately and in a mixed group- could facilitate this (Brooke-Sumner et al., 2024b). Whilst mental health NGOs in South Africa are facing significant funding challenges and require sustained support to do this work, this type of signposting and support is arguably less resource intensive- and therefore has greater potential for scalability and sustainability- compared to other options. However, disability grants did not meet all needs or provide financial security suggesting other approaches are also needed.

#### 2. Support to access existing savings groups and/or other income generation

Some participants had had positive experiences participating in stokvels, and that sometimes had more tangible impacts on financial security than disability grants. One option could be to use recovery groups, or similar psychosocial interventions, to discuss the benefits and risks of stokvels, along with practical advice on how to set up or join a group, but not to formally instigate the process as part of the intervention. This model would not be resource intensive to providers so theoretically would have good scalability and sustainability potential. However impoverished areas may be less likely to have existing groups. Moreover, existing community stokvels may be less likely to accept people living with SMHC, compared to caregivers, potentially further disempowering this group.

#### 3. Nesting savings groups within psychosocial interventions

Setting up savings groups targeting people with SMHC and caregivers could address their economic needs whilst circumventing problems with social exclusion. The model of nesting savings groups within recovery groups received broad support in our study. However our findings indicate that receipt of funds by either people with SMHC or caregiver alone could cause ruptures within families. As the financial security of people living with SMHC and caregivers is so highly linked, we suggest that the family may be the most appropriate unit of intervention delivery (Dijkstra et al., 2025). Nesting a savings element within a psychosocial intervention would also provide opportunity to actively address issues around autonomy of people living with SMHC and involvement in decision-making in a way that was sensitive to the social and cultural context. Our findings suggest that savings groups may work best when they arise organically and/or include people who know and trust each other. The positive social connections within PRIZE recovery groups that participants reported seem to offer powerful potential for successful group savings (Asher et al., 2024). This model would have moderate potential for scalability and sustainability; although input of capital would not be required, savings groups may require substantial support from providers to run successfully.

Whether sited within or outside a psychosocial intervention, there may be important barriers to the impact of the savings model. As the approach relies solely on the financial input of participants, and most families are living in poverty, the model only offers finite funds. Moreover some participants may have insufficient funds to contribute at all.

#### 4. Nesting income generation within psychosocial interventions

The wish of many participants was to start a small business, and this was seen as a having good potential to address financial insecurity. However, external input in form of equipment or capital was felt to be essential. A core component of the Basic Needs Model is to support livelihoods through savings and loans schemes and agricultural enterprises, alongside psychosocial support, community development and system strengthening (BasicNeeds, 2023). There is evidence from an uncontrolled evaluation that the Basic Needs model is cost effective and may have positive impacts on quality of life, functioning and income generation (Lund et al., 2013, de Menil et al., 2015). However, any model requiring ongoing external financial contributions would also have limited scalability and sustainability. This is an important consideration given the critical shortage of resources in most health systems, spanning both governmental and non-governmental agencies. Further, the approach of giving individual loans may not be appropriate given existing high levels of debt and stress caused by interactions with loan sharks in our study. Amongst highly impoverished South African populations without pre-existing mental health conditions, one study found some negative psychological effects of receiving ‘second look loans’ (that is, loans provided after a previous rejected application)(Fernald et al., 2008). However it is possible that providing loans to groups, rather than individuals, could offset some of these negative impacts. Yet it is also conceivable that provision of capital funds to savings groups may create disagreements within groups or increase the likelihood of being targeted by criminals. Multi and intersectoral approaches including working in partnership with organisations who are experienced in the design and delivery of savings groups and income generation would be paramount to ensure these concerns could be safely addressed using tested mechanisms (J. van Rensburg and Brooke-Sumner, 2023).

All four approaches would be enhanced by parallel social and public health interventions to reduce stigma and enhance social inclusion of people with SMHC (Asher et al., 2022), build social capital and reduce violence in neighbourhoods, including greater efforts by the criminal justice system (Rose-Clarke et al., 2020). These would offer a powerful way to address the underlying determinants we identified and help to make poverty alleviation interventions accessible and safe within neighbourhoods. We also note that this is not an exhaustive list of options but rather those that seem most feasible to provide as an adjuvant intervention to psychosocial support such as PRIZE recovery groups, and most relevant for low-resource settings. As such, whilst there is evidence that individual placement and support (in which people with SMHC receive support searching for competitive employment and mental health treatments concurrently) has positive impacts on vocational outcomes (Frederick and VanderWeele, 2019), limited employment opportunities in low-resource settings mean we do not propose this as a priority approach.

Future research to develop and evaluate poverty alleviation interventions for people with psychosis should use participatory approaches to unpick the complexities and realities of intervening in the economic ecosystem in low-resource and unstable communities. Hybrid effectiveness-implementation studies that investigate the impact for individuals alongside the practicalities, reach and sustainability of real-world implementation are needed. Furthermore evaluation designs that foreground the identification or measurement of negative unintended consequences of intervention would be essential. These might include dark logic models (Bonell et al., 2015), which aim to theorise harmful consequences of public health interventions, or causal loop diagrams (a technique to map interacting system dynamics, including positive and negative forces) (Jordans, 2025).

### Strengths and limitations

A strength of this study was the inclusion of individuals who had previously participated in PRIZE recovery groups. This meant they could comment meaningfully on how a poverty alleviation component could be incorporated in this type of psychosocial intervention. They also had experience of working in a group format with other people with SMHC and caregivers, which had revealed specific issues around power dynamics between caregivers and living with SMHC. Although we also included PRIZE participants who had not participated in recovery groups, we arguably focused on a narrow set of poverty alleviation interventions. For example, we did not explicitly seek participants views on conditional cash transfers or individual placement and support, which, if discussed, could have expanded our proposed options.

### Conclusion

Poverty alleviation interventions could positively impact on the wellbeing of people with SMHC and caregivers in South Africa as an adjunct to psychosocial interventions and psychiatric care. Approaches could include supporting access to social protection or existing savings groups, and nesting new savings groups or income generation initiatives into psychosocial interventions. Any approach would need to be carefully tailored to the needs of people with SMHC living in low-resource settings affected by high crime rates, such as promoting rights and equity, as well as incorporating robust mechanisms to ensure the safety of participants.

## Acknowledgements

The researchers thank Bharti Patel, Lameez Botha and the staff of the South African Federation for Mental Health and Indlela Mental Health Society for their support of PRIZE.

## Data availability

Due to the sensitive nature of the qualitative interviews and the risk of re-identification, data are available upon request from South African MRC Human Research Ethics Committee (coordinator Patricia Josias). Data will be shared with researchers who provide a methodologically sound research proposal and sign a data access agreement prohibiting re-identification and onward sharing.

## Notes

### Competing Interest Statement

The authors have declared no competing interest.

### Funding Statement

This work was supported by the South African Medical Research Council with funds received from the South African National Department of Health and the UK Medical Research Council, and with funds received from the UK Government’s Newton Fund. The funders had no role in the design of the study and collection, analysis, interpretation of data or in writing the manuscript.

### Author Declarations

The South African Medical Research Council Ethics committee (EC027-6/2021) gave ethical approval for the study

